# Evaluating ReFresh, a Web-Based Alternative to Suspension for Adolescent E-cigarette Use

**DOI:** 10.64898/2026.10.05.26364747

**Authors:** Leslie D. Litsky, Brittanie Moore

## Abstract

Electronic cigarettes are the most commonly used tobacco product among U.S. adolescents. Schools most often respond to student e-cigarette use with exclusionary discipline such as suspension, which removes students from school and does not address potential underlying nicotine dependence. Web-based alternatives to suspension offer a way to keep students in school while providing cessation-focused support, but evidence on their effects is limited. This study examined changes in e-cigarette-related attitudes and motivation among adolescents who completed ReFresh, a self-paced, web-based alternative-to-suspension program completed in approximately 3 hours. Using a single-group pre-post design, we analyzed data from 173 middle and high school students (grades 6 to 12; mean age 15.0) referred after being caught using e-cigarettes on school grounds across 24 delivery sites (e.g., schools, afterschool programs, and health centers). The program comprises five interactive online lessons grounded in the 5 A’s clinical framework (Ask, Advise, Assess, Assist, Arrange), motivational interviewing, and cognitive and behavioral skills training. Changes were analyzed with non-parametric tests. After completing ReFresh, participants reported greater motivation to quit (χ2 (6) = 22.79,*OR* = 3.80, 95% CI [2.2,8.8], *p* < .001) and rated quitting as more important (*V* = 3037.50,95% CI [1.00, 2.00], *p* < .001, *r* = 0.2). They also viewed e-cigarette use as more harmful and more risky (*V* = 8137.00, 95% CI [0.50, 0.63, *p* < .001, *r* = 0.62; *V* = 7734.50, 95% CI [0.50,0.80), *p* < .001, *r* = 0.65)), with the largest shifts in perceived harm relative to cigarettes and perceived risk of mental health problems. Most students reported comfort and confidence seeking help for e-cigarette use or mental health concerns, ranging from 71.10% of students agreeing they felt “More confident asking a trusted adult for help at school” to 80.50% of students agreeing they felt “More confident encouraging a peer to seek help”. This study suggests that a brief, web-based alternative to suspension may shift the attitudes and motivation that precede quitting while keeping students in school, offering schools a non-exclusionary way to respond to e-cigarette use. Controlled studies measuring actual e-cigarette behavior are needed to confirm whether these programs reduce use.

## INTRODUCTION

Electronic cigarettes (e-cigarettes) are the most commonly used tobacco product among U.S. adolescents. (1) In 2024, 5.9% of middle and high school students reported current e-cigarette use, including 7.8% of high school students. (2) While overall prevalence has fallen from its 2019 peak, those who continue to vape often do so heavily. Among students who currently vape, 38.4% report frequent use and 26.3% report daily use. (1) Sustained use at these levels makes adolescent e-cigarette use a continuing concern for adolescent health. Given that much of this use occurs during the school day and interferes with daily operations, schools are often forced to address the issue. (3)

E-cigarette use is not a benign behavior. Most e-cigarettes deliver nicotine, which is highly addictive and can harm the developing adolescent brain, affecting attention, learning, mood, and impulse control. (4,5) Adolescent e-cigarette use is also consistently associated with poor mental health. Youth who vape report higher rates of depression and anxiety than those who do not. (6) This relationship appears bidirectional, as many adolescents report vaping to cope with stress or anxiety, while nicotine addiction and withdrawal can in turn worsen those symptoms, reinforcing a cycle of dependence. (6)

Schools most often treat e-cigarette use as a disciplinary infraction rather than a health problem. In a California survey of nearly 8,000 school staff, the most common responses to students caught using e-cigarettes were exclusionary, including detention, suspension, expulsion, and referral to law enforcement. (7) The American Academy of Pediatrics classifies exclusionary discipline as a threat to child health and recommends reserving it for the most serious cases. This is particularly important for Black, American Indian/Alaska Native, male, and LGBTQ+ students as they are suspended and expelled at disproportionately high rates overall. (8) Removing a student from school also carries documented academic harm. A meta-analysis of 34 studies found that suspension is inversely related to academic achievement and positively related to dropout. (9) Even a single suspension is associated with roughly double the risk of dropping out of high school. (10) Another study extended these findings by analyzing a redistricting boundary change that reassigned students across schools. Students assigned to higher-suspension schools had lower educational attainment and higher rates of adult arrest and incarceration. (11) These harms are not limited to suspended students. Schoolwide, higher suspension rates are also associated with lower academic achievement even among peers who were never disciplined. (12) Suspension also reduces school connectedness, a protective factor against adolescent risk behaviors including substance use, which is counterproductive when the infraction is a health-risk behavior.(13) Discipline also does nothing about the potential nicotine dependence itself. Federal clinical guidance recommends counseling and behavioral cessation support for adolescents trying to quit, neither of which a suspension provides. (14) This gap between how schools respond and what adolescents would benefit from has prompted a need for alternatives.

In response to these harms, many schools have adopted alternatives that address behavior while keeping students in school, most commonly restorative practices, which emphasize accountability, relationship repair, and community rather than removal. (15) A systematic review of 37 such interventions found they can reduce exclusionary discipline in the short term. (16) A randomized controlled trial in the Pittsburgh Public Schools found that a whole-school restorative program reduced suspension rates and improved teacher-rated school climate, with larger reductions among elementary, Black, low-income, and female students. (17) The same trial found mixed-to-negative effects on school-level academic outcomes and was highly dependent on implementation support. These results show that schools can reduce exclusion without removing students, though the effects of the interventions vary.

Schools have developed alternative-to-suspension programs specifically for e-cigarette use. These programs typically replace removal with required participation in education, goal-setting, and cessation support delivered by school staff. (18) However, evidence for their effectiveness remains limited. Reviews report mixed effects on adolescent tobacco-related beliefs and behavior, and there are few rigorous cessation trials in this population. The ones that exist have shown limited abstinence effects. (18,19) Most evaluations of school-based e-cigarette programs have focused on preventing initiation. (18) Alternative-to-suspension programs are less studied, especially brief, web-based ones and their effect on students’ attitudes and motivation to quit is largely unknown. One factor shaping whether such programs work is how well they match what adolescents themselves say they need. Research on youth preferences indicates that adolescents want cessation programs to be confidential and to teach practical skills such as stress management, alongside clear information on the health effects of e-cigarettes.(20) Most of these programs also rely on in-person, staff-delivered formats, which limit how widely and consistently they can be offered. They often depend on trained staff, scheduling, and school resources that vary by site. Digital tools could address these constraints by delivering consistent content at scale and reaching students across many schools regardless of local capacity, but such programs remain understudied. Yet it remains unclear how brief, web-based programs can deliver these features and shift students’ thinking about e-cigarettes.

ReFresh is a web-based alternative-to-suspension program for middle and high school students found using nicotine e-cigarettes in school. Delivered as a self-paced online session with embedded quizzes, it can be completed in one or multiple settings. ReFresh serves as an educational component of a school’s overall response to vaping, providing a non-punitive alternative designed to eliminate or reduce suspension days. Assigned as an immediate educational opportunity, the program delivers targeted behavioral support which keeps the student connected to the academic environment. School staff often prefer an opportunity that can be applied promptly after an e-cigarette incident, both to maintain consistent policy enforcement and to intervene while the behavior is recent. (7) This study addresses the gap in evidence about whether a low-intensity, scalable program, such as ReFresh, can shift the attitudes and motivation that precede quitting. (21)

This study evaluated changes in e-cigarette-related attitudes and motivation among adolescents who completed ReFresh under routine school conditions. Using a single-group pre-post design, we examined whether students’ motivation to quit, perceived importance of quitting, confidence in quitting, and perceptions of e-cigarette harms changed before and after the program. We also described students’ post-program attitudes toward seeking help for e-cigarette use and mental health concerns.

## METHODS

### Program Overview

ReFresh is a web-based, self-paced alternative-to-suspension program for middle and high school students (grades 6 to 12) who have been identified with nicotine e-cigarette use or possession on school grounds. ReFresh was developed by Playbl, a spinout organization from the play2PREVENT Lab at Yale University School of Medicine. It is used by schools in multiple ways. It was used to either replace suspension or reduce the number of suspension days. This keeps students in school while delivering cessation-focused education and behavioral support. The curriculum consists of five sequential lessons, each with three to six topics, delivered through interactive videos with embedded reflection prompts, quiz items, and worksheet activities. It requires approximately three hours of instructional time, typically completed across one to three sessions.

ReFresh is hosted on a custom Playbl web platform and completed on internet-connected devices. Students log in through a web portal using individual accounts created before they begin the program, most often via school-provided Google or Microsoft single sign-on. The platform enforces linear progression. Students cannot fast-forward or skip ahead, and each topic unlocks only after they complete the previous one. Content is delivered through interactive videos with full voiceover narration, peer narratives from youth presenters, and a recurring cast of animated adolescent characters whose storylines span the curriculum.

The five lessons follow the program’s progression from awareness to skill building and goal setting. Lesson 1, Why Do I Vape?, builds self-awareness through personal stories, trigger reflection, and education on nicotine addiction. Lesson 2, Understanding and Responding to My Thoughts and Feelings, teaches cognitive reframing, emotional regulation, and decision making skills. Lesson 3, What Happens to My Mind and Body When I Vape, covers the physical and mental health effects of nicotine, including organ health and sleep. Lesson 4, Your Body’s Reset, provides facts about withdrawal, habit replacement and craving management tools. Lesson 5, My Next Play for a Healthier Me, builds help seeking skills, connects students to resources, and guides creation of a personalized Vaping Quit Plan. See Supplemental File for more information on lessons.

ReFresh is grounded in established frameworks for behavior change and cessation. Content is structured around the clinical 5 A’s framework (Ask, Advise, Assess, Assist, Arrange) for tobacco and e-cigarette cessation, and draws on motivational interviewing, cognitive behavioral therapy, dialectical behavior therapy, and mindfulness.(22–25) Beyond health education, the program teaches self-regulation skills such as goal-setting and emotional control.

### Pilot Testing

From October 2024 to March 2025, ReFresh underwent a development and pilot testing phase in which the curriculum was reviewed in stages by a clinical expert now at Chobanian & Avedisian School of Medicine, school staff, and adolescent testers. Seven school administrators, social workers, and disciplinary staff evaluated the program structure through focus groups to assess feasibility within existing disciplinary models. Their feedback shaped several design features, including full voiceover narration, linear navigation that prevents skipping, and exportable student journals and quit plans for staff follow-up. Three adolescent testers reviewed the program’s usability and pacing, noting that interactive exercises fostered reflection, coverage extended to mental health and sleep impacts, and the empowering tone reinforced their capacity to quit. These rounds of feedback informed platform and content revisions before ReFresh launched in March 2025.

#### Measures

Pre- and post-assessments were administered within the ReFresh platform and linked at the individual student level. They were non-graded and confidential. The pre-assessment included four demographic items (age, sex, race/ethnicity, grade) and a 13-item core scale measuring motivation to quit, importance and confidence, perceived harm, perceived risk, and perceived dangerousness. Motivation was measured with a single item adapted from the Motivation to Stop Scale, rated on a 7-point ordered scale. (26) Importance of quitting and confidence to quit were each measured on 0 to 10 scales adapted from the Motivational Interviewing rulers.(27) Two subscales assessed perceptions of e-cigarette use on 1 to 5 Likert scales: perceived harm (4 items: harm to overall health, harm to the lungs, harm relative to cigarettes, and harm from occasional use; α = .81), perceived risk (5 items: likelihood of serious health problems, nicotine addiction, lung disease, heart disease, and mental health problems; α = .85). Perceived dangerousness was assessed by participants rating their agreement with the statement “Vaping is dangerous for your health” on a 1 to 5 Likert scale. Items were adapted from established youth tobacco surveillance instruments, including the National Youth Tobacco Survey(28), Monitoring the Future(29), and the Michigan Profile for Healthy Youth.(30) Subscale scores were computed as the mean of their items. Higher scores indicated a more health-protective direction on all items.

The post-assessment added six help-seeking items (1 to 5 Likert) measuring comfort and confidence in seeking help for e-cigarette use or mental health concerns, and three open-ended prompts asking what students liked, disliked, and would change.

#### Procedures

Implementation began in March 2025 across more than 50 schools. As of July 2026, 173 students had completed the program across 24 delivery locations, including schools, afterschool programs, and health centers. The remaining sites had not yet enrolled or completed a participant within the study window. Delivery occurred in three settings, determined by the contracting agency: school-based settings (63.6%, n = 110), including in-school suspension rooms, supervised school locations, alternative schools, and supervised take-home assignments; community-based organizations (24.9%, n = 43), such as after school or weekend disciplinary programs; and health department extension sites (11.6%, n = 20) managing youth disciplinary referrals. Students completed the demographic questions and the motivation, importance, confidence, harm, risk, and dangerousness measures immediately before completing the ReFresh program. After the ReFresh program, students completed the motivation, importance, confidence, harm, risk, and dangerousness measures again along with the added help-seeking and open-ended feedback questions.

#### Ethics Review

This study was determined to be exempt by the Heartland Institutional Review Board Project No. 042126-1437, under exemption category #, 45 CFR 46.104(d)(1) - 46.104(d)(8)], Secondary Research Uses of Data/Biospecimens (46.104(d)(4)).

#### Statistical Analysis Plan

This study used a single-group pre-post design to evaluate change in e-cigarette-related outcomes among students who completed ReFresh. Each participant served as their own comparison. For items administered both before and after the program, changes were assessed with paired (within-subject) tests. Help-seeking items, administered only post-program, were summarized descriptively without comparison.

Non-parametric tests were used to assess ordinal and non-normal data. Normality and symmetry of difference scores were examined to select the appropriate test for each item. Difference scores were non-normal for all measures except the harm and dangerousness subscales. For motivation, confidence, harm, and dangerousness, difference scores were leptokurtic but symmetric about the median, so Wilcoxon signed-rank tests were used. For the importance of quitting, difference scores were leptokurtic with heavy positive skew, so a sign test was used.

## RESULTS

### Participants

Participants were 173 students in grades 6 through 12 who were caught using or possessing nicotine e-cigarette products on school grounds and referred to ReFresh by staff as an alternative to suspension. No eligibility criteria were applied beyond staff referral following a disciplinary infraction and grade level. Ages ranged from 11 to 18, with one participant aged 19 or older (*M*= 15.04, *SD* = 1.76). High school students made up 72.1% of the sample. The sample was 58.2% male and 41.2% female. Self-reported race and ethnicity were 57.0% White, 17.7% Hispanic/Latino, 13.3% mixed race, and 6.3% Black. Completers were defined as students who finished all five lessons and the post-assessment. This analysis only included data from those who completed ReFresh.

Of the 231 students referred to ReFresh during the study window, 173 completed all five lessons and the post-assessment and were included in the analysis. Sample characteristics are described in the Participants section. Analyses paired each student’s pre- and post-responses. The number of complete pairs varied by item due to missing responses, ranging from 171 to 173.

#### Motivation to Quit

Participants reported greater motivation to quit e-cigarettes after completing ReFresh. A Wilcoxon signed-rank test showed a significant but small increase (*V* = 2504.00, 95% CI [ 0.50,1.50], *p* < .001, *r* = 0.28), with the median rising from 5 to 6 and the mean from 4.96 (*SD* = 2.03) to 5.49 (*SD* = 1.78). Most participants showed no change (90, 52.63%), and 26 (15.20%) scored lower at post-test.

Because the motivation item used seven closely worded response options, we examined these individual shifts. Most declines were movements between adjacent middle categories with similar meaning and no participant moved from wanting to quit to not wanting to quit. In an exploratory analysis, we collapsed the item into four categories corresponding to Transtheoretical Model stages (see Table 2 for the mapping). Under this coding, only 13 participants (7.6%) declined, and McNemar’-Bowker test confirmed an overall shift toward higher motivation (χ2 (6) = 22.79,*OR* = 3.80, 95% CI [2.2,8.8], *p* < .001).

**Table 1.** Participant Characteristics (N = 173)

| Characteristic | n | % |
| --- | --- | --- |
| *Grade level* |  |  |
| High school (9–12) | 125 | 72.1 |
| Middle school (6–8) | 48 | 27.9 |
| *Age* |  |  |
| Mean (SD) | 15.04 (1.76) | — |
| Range | 11–19 | — |
| *Sex* |  |  |
| Male | 101 | 58.2 |
| Female | 71 | 41.2 |
| Not reported | 1 | 0.6 |
| *Race/ethnicity* |  |  |
| White | 99 | 57.0 |
| Hispanic/Latino | 31 | 17.7 |
| Mixed race | 23 | 13.3 |
| Black | 11 | 6.3 |
| Not reported/other | 9 | 5.7 |

**Table 2.**
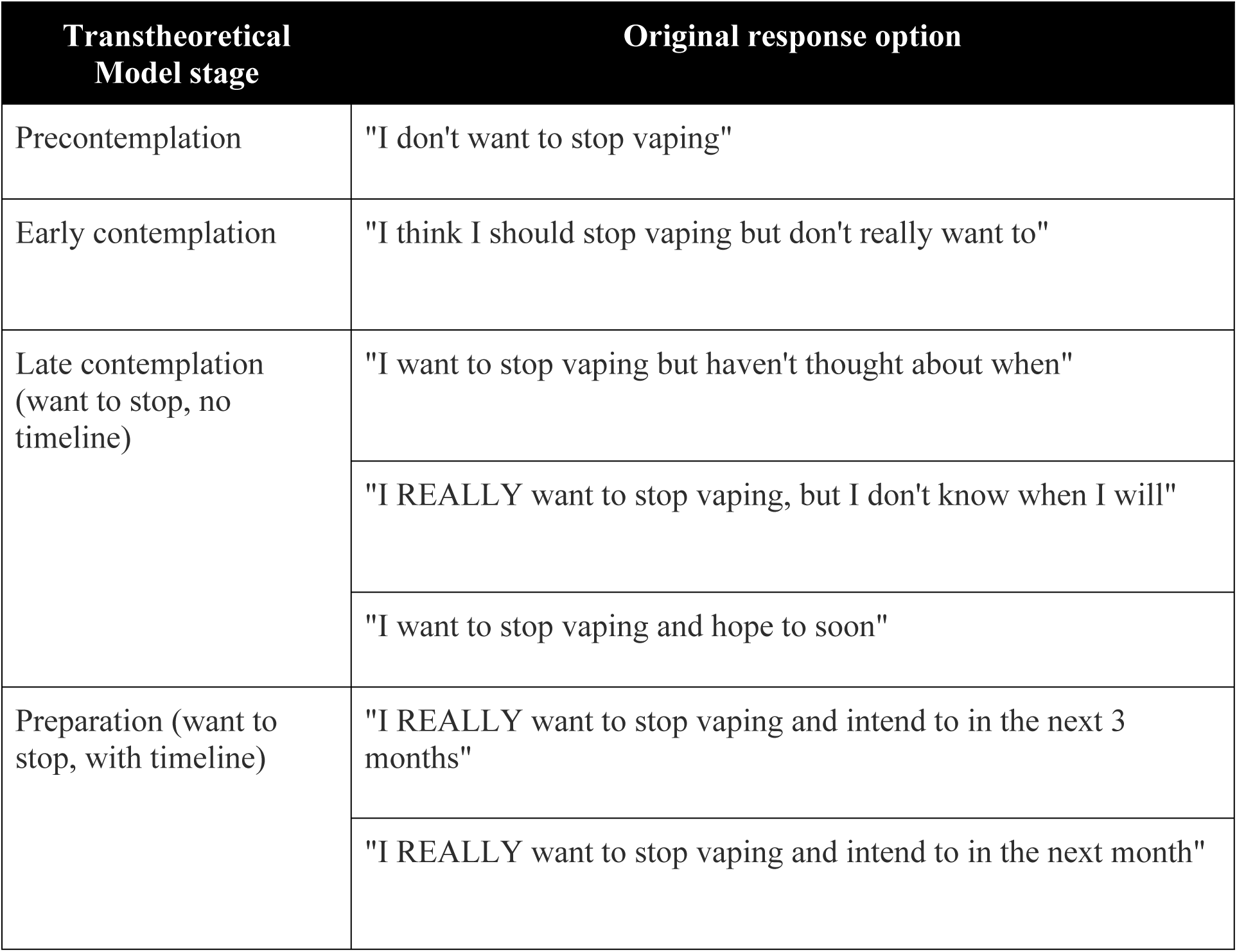

### Importance and Confidence

Alongside motivation, participants’ sense of how important it was to quit and their confidence in their ability to quit also increased. A sign test showed participants rated quitting as more important after the program (*S* = 150,95% CI [3.00,3.00], *p* < .001, Cohen’s *g* = 0.40). The median rose from 8 to 10 and the mean from 7.17 (*SD* = 3.36) to 8.10 (*SD* = 2.91), with 70.2% of participants showing a positive change, 2.9% showing no change, and 9.4% showing a decrease. Confidence to quit showed a smaller shift. A Wilcoxon signed-rank test indicated a significant increase (*V* = 3037.50,95% CI [1.00, 2.00], *p* < .001, *r* = 0.28), though the median remained at 9 and the mean rose only from 7.53 (*SD* = 3.07) to 8.33 (*SD* = 2.21). Nearly half of participants showed no change (46.8%), while more increased (33.9%) than decreased (17.0%), a pattern consistent with a ceiling effect given the high baseline confidence.

### Perceived Harm and Risk

Participants rated e-cigarette use as more harmful and more risky after completing ReFresh. On the perceived harm subscale, a Wilcoxon signed-rank test showed a significant increase (*V* = 8137.00, 95% CI [0.50, 0.63, *p* < .001, *r* = 0.62), with the median rising from 3.75 to 4.25 and the mean from 3.79 (*SD* = 0.75) to 4.17 (*SD* = 0.60). The perceived risk subscale increased similarly (*V* = 7734.50, 95% CI [0.50,0.80), *p* < .001, *r* = 0.65), with the median rising from 3.80 to 4.60 and the mean from 3.79 (*SD* = 0.94) to 4.33 (*SD* = 0.71). Perceived dangerousness also increased (*V* = 2463.50, 95% CI [1.00,1.00], *p* < .001, *r* = 0.41), with the median rising from 4 to 5 and the mean from 4.16 (*SD* = 0.94) to 4.52 (*SD* = 0.71).

Although the harm and risk subscales had very good internal consistency, we examined item-level change to see whether shifts differed by type of health concern. The largest increases were in beliefs that e-cigarette use is more harmful to overall health than cigarettes (*M* = 3.36, *SD* = 1.12, to 4.34, *SD* = 0.91) and that e-cigarette use raises the risk of anxiety and other mental health problems (*M* = 3.47, *SD* = 1.34, to 4.28, *SD* = 0.92). The smallest increases were in perceived harm from occasional, non-daily use and in perceived risk of lung disease. Item-level means and standard deviations are shown in Table 3.

**Table 3:** Changes in Motivation and Risk Perceptions Before and After ReFresh (N = 173)

| Outcome | Pre M<br>(SD) | Post M<br>(SD) | Pre<br>Mdn | Post<br>Mdn | Test | p |
| --- | --- | --- | --- | --- | --- | --- |
| Motivation to quit | 4.96 (2.03) | 5.49 (1.78) | 5 | 6 | V = 817.00 | < .001 |
| Importance of quitting | 7.17 (3.36) | 8.10 (2.91) | 8 | 10 | Sign test | < .001 |
| Confidence to quit | 7.53 (3.07) | 8.33 (2.21) | 9 | 9 | V = 1148.50 | < .001 |
| Perceived harm (subscale) | 3.79 (0.75) | 4.17 (0.60) | 3.75 | 4.25 | V = 1043.00 | < .001 |
| Perceived risk (subscale) | 3.79 (0.94) | 4.33 (0.71) | 3.80 | 4.60 | V = 780.50 | < .001 |
| Perceived dangerousness | 4.16 (0.94) | 4.52 (0.75) | 4 | 5 | V = 539.50 | < .001 |

### Help-Seeking (post-only)

At the end of the program, participants responded to six items about seeking help for e-cigarette use or mental health concerns. Because these items were administered only after ReFresh, no pre-post comparison was possible. Across all six items, most participants agreed or strongly agreed: between 71% and 80% endorsed each statement, including feeling comfortable asking for help at school, feeling confident asking a trusted adult for help inside and outside of school, being more likely to seek help in both settings, and feeling confident encouraging a peer to seek help (80.5% agreed or strongly agreed). Item-level percentages are shown in Table 4.

**Table 4:** Post-Program Help-Seeking Attitudes (N = 173)

| Help-seeking item | Strongly agree (%) | Agree (%) | Neutral (%) | Disagree (%) | Strongly disagree (%) |
| --- | --- | --- | --- | --- | --- |
| Felt more comfortable asking for help at school | 46.82 | 26.59 | 17.92 | 4.62 | 4.05 |
| More confident asking a trusted adult for help at school | 47.98 | 23.12 | 19.08 | 6.36 | 4.47 |
| More confident asking a trusted adult for help outside school | 50.87 | 25.43 | 17.34 | 3.47 | 2.89 |
| More likely to seek help at school | 45.09 | 28.32 | 17.34 | 6.36 | 2.89 |
| More likely to seek help outside school | 48.55 | 27.17 | 19.08 | 4.05 | 1.16 |
| More confident encouraging a peer to seek help | 52.02 | 28.48 | 13.87 | 2.89 | 1.73 |

## DISCUSSION

This study examined changes in e-cigarette-related attitudes, perceived harm and risk, motivation to quit, and help-seeking confidence among adolescents who completed ReFresh, a brief web-based alternative-to-suspension program. After completing the program, participants reported greater motivation to quit, rated quitting as more important, and viewed e-cigarettes as more harmful, riskier, and more dangerous than before completing the program. Most participants also expressed confidence in seeking help for e-cigarette use or mental health concerns. The most relevant changes were in the motivational precursors to quitting. Participants reported greater motivation and rated quitting as more important after the program, consistent with the program’s grounding in the 5 A’s framework and motivational interviewing, which target readiness and personal relevance rather than information alone. Results suggest ReFresh moved students along the early steps toward quitting.

Participants also came to see e-cigarette use as more harmful and more risky. The largest change in the harm items was in how harmful students judged e-cigarettes to be, relative to cigarettes. This is notable because adolescents commonly underestimate the harms of e-cigarettes and perceive them as safer than combustible cigarettes, a belief associated with greater willingness to try and continue e-cigarette use. (31,32) Among the risk items, the largest change was in the perceived likelihood that e-cigarettes contribute to anxiety and other mental health problems. This pattern aligns with what adolescents themselves say they want from cessation programming, including an emphasis on the link between nicotine, stress, and emotional regulation rather than physical health warnings alone.(20) It is notable that the mental health item moved most given that stress management is among the skills youth most often request.

After completing ReFresh, most participants expressed comfort and confidence in seeking help for e-cigarette use or mental health concerns, and a majority reported confidence encouraging a peer to do the same. Because these items were measured only after the program, they describe post-program attitudes rather than change, but they align with the program’s aim of connecting students to support rather than removing them from it. This is where an alternative-to-suspension approach departs most clearly from exclusionary discipline. Suspension severs the school connectedness that protects adolescents against health-risk behaviors (33,34), whereas a program like ReFresh can turn the same disciplinary moment toward help-seeking and follow-up. This matters because adolescents who vape often do not seek help, and willingness to approach a trusted adult is itself a protective factor.(35) The exportable quit plans offer one mechanism for that follow-up, giving counselors and social workers a concrete starting point for continued support. Whether such follow-up occurs or improves outcomes was not measured here and should be an area for future research. Prior reviews have found inconsistent effects on adolescent tobacco-related beliefs and behavior, and rigorous evaluations of alternative-to-suspension programs for e-cigarettes specifically remain scarce.(18,36) By documenting change across attitudes, risk perceptions, and motivation, this study suggests that a brief, web-based program can influence the precursors to quitting.

Beyond its content, ReFresh’s format may shape its usefulness to schools. It is brief, self-paced, and delivered entirely online, addressing two recurring problems in school-based interventions. First, restorative and alternative-to-suspension approaches often depend heavily on staff training and consistent delivery, and their effects vary with implementation fidelity.(17,37,38) As a standardized digital program, ReFresh delivers the same content to every student regardless of a school’s capacity, staffing, or expertise, removing a common source of variation. Second, the format aligns with what adolescents say they want from cessation programming: privacy and self-paced progression.(20) Completing a program alone on a device, rather than in a group or a face-to-face meeting, may lower the embarrassment or stigma that discourages some students from engaging. These same features are both a strength and a limitation. A self-paced digital program cannot offer the responsive, relational support of a skilled counselor, and it remains unknown whether a brief online session produces durable change, as opposed to the immediate attitudinal shifts observed here. Future studies could examine whether a program like ReFresh works best on its own or as a complement to counselor-led or in-person support, combining the consistency of digital delivery with the responsiveness of a person.

This evaluation has several limitations. First, the single group pre-post design lacked a control group. Although each participant served as their own comparison, concurrent influences such as the disciplinary event itself, maturation, or social desirability could also account for the observed changes. Second, the analytic sample included only students who completed all five lessons and the post-assessment. Those who did not complete the program were not captured, and completers may differ from non-completers in ways that affect the results. Third, all outcomes were self-reported within the platform, and although responses were confidential and non-graded, self-report in a disciplinary context may be subject to social desirability bias. Fourth, the outcomes measured attitudes, perceived harm, and motivation, not actual e-cigarette behavior. The study does not establish whether the program reduced e-cigarette use or produced lasting change. Outcomes were also assessed immediately after completion, so durability is unknown, and the help-seeking items were administered only after the program, precluding pre-post comparison. Additionally, this was a secondary analysis of data collected during routine program implementation rather than a study designed to test the program’s effects. Finally, motivation, importance, and confidence were each measured with a single item, which limits their reliability. Despite these limitations, findings suggest that brief, web-based alternative-to-suspension programs may offer schools a non-punitive way to respond to student e-cigarette use while keeping students in class.

## Conclusion

Among adolescents referred for e-cigarette use, completing ReFresh was associated with increased motivation to quit, greater perceived importance of quitting, and stronger perceptions of harm and risk. These findings add to a growing effort to replace exclusionary discipline with responses that treat e-cigarettes as a health issue rather than a rule violation. As schools search for ways to address student e-cigarette use without removing students from the classroom, brief, web-based alternatives to suspension offer a promising, scalable option that keeps students in school, connects them to support, and begins to shift the attitudes and motivation that precede quitting. Approaches like ReFresh appear promising in helping schools respond to e-cigarettes in ways that keep students learning and connected to support.

## Data Availability

No data was generated by this study. An existing data set was used but is not available publicly for inquiries. Data can be provided upon reasonable request.

## Acknowledgments

We gratefully acknowledge the following individuals for their essential contributions to the development and evaluation of ReFresh:

• **Zoe Thanawala**, ORISE Fellow at the Centers for Disease Control and Prevention (2021–2023), for her foundational contributions to the original product design, theoretical framework, literature review, and curriculum content development.

• **Sarah Bagley, MD**, Associate Professor of Medicine and Pediatrics at Chobanian & Avedisian School of Medicine, for her expert review of health messaging and medical accuracy.

## Notes

### Competing Interest Statement

The authors have declared no competing interest.

